# Clinical prediction rule for SARS-CoV-2 infection from 116 U.S. emergency departments

**DOI:** 10.1101/2021.01.20.21249656

**Authors:** Jeffrey A. Kline, Carlos A. Camargo, D. Mark Courtney, Christopher Kabrhel, Kristen E. Nordenholz, Thomas Aufderheide, Joshua Baugh, David Beiser, Christopher Bennett, Joseph Bledsoe, Edward Castillo, Makini Chisholm-Straker, Elizabeth Goldberg, Hans House, Stacey House, Timothy Jang, Chris Kabrhel, Stephen Lim, Troy Madsen, Danielle McCarthy, Andrew Meltzer, Stephen Moore, Craig Newgard, Justine Pagenhardt, Katherine L. Pettit, Michael Pulia, Michael Puskarich, Lauren Southerland, Scott Sparks, Danielle Turner-Lawrence, Marie Vrablik, Alfred Wang, Anthony Weekes, Lauren Westafer, John Wilburn

## Abstract

**Objectives:** Accurate and reliable criteria to rapidly estimate the probability of infection with the novel coronavirus-2 that causes the severe acute respiratory syndrome (SARS-CoV-2) and associated disease (COVID-19) remain an urgent unmet need, especially in emergency care. The objective was to derive and validate a clinical prediction rule for SARS-CoV-2 infection that uses simple criteria widely available at the point of care.

**Methods:** Data came from the Registry data from the national REgistry of suspected COVID-19 in EmeRgency care (RECOVER network) comprising 116 hospitals from 25 states in the US. Clinical predictors and 30-day outcomes were abstracted from medical records of 19,850 emergency department (ED) patients tested for SARS-CoV-2. The criterion standard for diagnosis of SARS-CoV-2 required a positive molecular test from a swabbed sample or positive antibody testing within 30 days. The prediction rule was derived from a 50% random sample (n=9,925) using unadjusted analysis of 107 candidate variables as a screening step, followed by stepwise forward logistic regression on 72 variables.

**Results:** Multivariable regression yielded a 13-variable score, which was simplified to 13-point rule: +1 point each for age>50 years, measured temperature>37.5°C, oxygen saturation<95%, Black race, Hispanic or Latino ethnicity, household contact with known or suspected COVID-19, patient reported history of dry cough, anosmia/dysgeusia, myalgias or fever; and -1 point each for White race, no direct contact with infected person, or smoking. In the validation sample (n=9,975), the score produced an area under the receiver operating character curve of 0.80 (95% CI: 0.79-0.81), and this level of accuracy was retained across patients enrolled from the early spring to summer of 2020. In the simplified rule, a score of zero produced a sensitivity of 95.6% (94.8-96.3%), specificity of 20.0% (19.0-21.0%), likelihood ratio negative of 0.22 (0.19-0.26). Increasing points on the simplified rule predicted higher probability of infection (e.g., >75% probability with +5 or more points).

**Conclusion:** Criteria that are available at the point of care can accurately predict the probability of SARS-CoV-2 infection. These criteria could assist with decision about isolation and testing at high throughput checkpoints.

**Key points:** *Question:* Can clinical criteria, derived solely from interview and vital signs accurately estimate the probability of infection from the novel coronavirus (SARS-CoV-2) that causes COVID-19?

*Findings:* From derivation sample (n=9,925), we derived a set of 13 clinical criteria that produced an area under the receiver operating characteristic curve of 0.80 (0.79-0.81) in a validation sample (n=9,925). At a score of zero, the simplified version of the criteria produced sensitivity of 95.6% (94.8 to 96.3%), and specificity of 20.0% (19.0 to 21.0%).

*Meaning:* Clinical criteria can estimate the probability of SARS-CoV-2 infection.

## BACKGROUND

The ability to rapidly estimate the probability of infection with the novel coronovirus-2 that causes severe acute respiratory syndrome (SARS-CoV-2) remains a formidable problem. The protean clinical picture of SARS-CoV-2 infection confounds its prediction. For example, the disease syndrome that SARS-Cov-2 produces--recognized as COVID-19--can manifest a wide range of nasopharyngeal, respiratory, and gastrointestinal symptoms, and a substantial minority of patients who carry SARS-CoV-2 manifest no symptoms at the time of testing.^1,2^ Asymptomatic patients can manifest nasopharyngeal viral loads, and shedding capacity similarly to symptomatic, infected persons.^3,4^ Factors limiting our current knowledge include the lack of systematically collected data from a large, unbiased, geographically diverse samples of patients, and problems associated with limited availability of molecular diagnostic tests and assays, long turnaround time and low diagnostic accuracy.^3,5-7^ The need for rapid exclusion without molecular testing arises daily in health care clinics, outpatient treatment facilities, at the point of intake for homeless shelters, judicial centers for incarceration, and extended care facilities. This need is particularly urgent in the emergency department (ED), which represents the largest interface between the general public and unscheduled medical care. In 2016, the >5000 US EDs had approximately 145 million patients.^8^ Additionally, because the ED interconnects with both outpatient and inpatient medical care, the critical question of SARS-CoV-2 infection status affects decisions to admit or discharge the patient, return to work, need for home isolation, and the location of hospital admission. These questions become more complicated for patients without access to basic medical care, and those experiencing serious mental illness, substance use disorders, and homelessness.

To address these needs, the authors created the <u>RE</u>gistry of suspected <u>COV</u>ID-19 in <u>E</u>me<u>R</u>gency care (RECOVER), a national network to capture data from patients tested for SARS-CoV-2 and evaluated in the ED.^9^ This report addresses the primary goal of the initial network-wide registry, which was to create a quantitative pretest probability scoring system (putatively named the COVID-19 Rule Out Criteria [CORC score]) to predict a positive SARS-CoV-2 test, and relatedly the derivation and internal validation of a simplified version that can serve as a prediction rule to identify those at very low probability of disease (the CORC rule). The intent of the rule was to function similarly to the Wells pretest probability scoring criteria and Pulmonary Embolism Rule out Criteria (PERC rule) for acute pulmonary embolism, respectively, except the diagnostic target was SARS-CoV-2.^10,11^

## METHODS

The RECOVERY network has resulted from the collaboration of 45 emergency medicine clinician-investigators from unique medical centers in 27 US states. Most of the 45 medical centers participating are the flagships of hospital networks that include community and academic centers. Information about these sites, and the methods of the initial registry, are available elsewhere.^9^ The primary objective of the registry was to obtain a large sample of ED patients with suspected SARS-CoV-2 and who had a molecular test performed in the ED as part of their usual care. The design, collection, recording and analysis of data for this report were conducted in accordance with the transparency in reporting of a multivariable prediction model for individual diagnosis and prognosis (TRIPOD) criteria.^12^ The RECOVER registry protocol was reviewed by the institutional review boards (IRBs) at all sites; 42 IRBs provided an exemption from human subjects designation, whereas three IRBs provided approval with waiver of informed consent.

Briefly, eligibility for enrollment required that a molecular diagnostic test was ordered and performed in the ED setting with suspicion of possible SARS-CoV-2 infection, or COVID-19 disease.^9^ Patients could only be enrolled once. Otherwise, there were no age or symptom-based exclusions; however, the guidance was provided to exclude patients where the test was clearly done for automated, administrative purposes in the absence of any clinical suspicion for infection. One example for exclusion was patients without suspected infection but who had swab testing performed in the ED done only to comply with a hospital screening policy for admissions or pre-operative testing. All sites were contracted to abstract charts from at least 500 patients.

Data were collected from the electronic medical record, using a combination of electronic download for routinely collected, coded variables (e.g., age, vital signs and laboratory values), supplemented by chart review by research personnel, using methods previously described.^9^ Each data form included 204 questions resulting in 360 answers, because many questions allowed multiple answers. Data were archived in the REDCap® system, with electronic programming to ensure completion of mandatory fields and sensible ranges for parametric data. Training of data abstractors was done via teleconference with the principal investigator (JAK) and program manager (KLP), supplemented by an extensive guidance document and field notes present in the REDCap® system, visible to the person doing enrollment. This analysis was pre-planned as the first manuscript from the RECOVER network.

To generate a comprehensive pool of independent predictors for a quantitative pretest probability model, as well as harmonization with other data, we recorded 28 symptoms, including all symptoms from the Clinical Characterization Protocol from the World Health Organization-supported International Severe Acute Respiratory and Emerging Infection Consortium (ISARIC).^13^ The form also recorded 14 contact exposure risks, ranging from no known exposure, to constant exposure to a household contact with COVID-19. We anticipated that many patients would have multiple ED visits prior to testing, especially for atypical presentations, and the goal was to collect patient data from the earliest medical presentation. Accordingly, the symptoms and contact risks, together with the vital signs (body temperature, heart rate, respiratory rate, systolic and diastolic blood pressure and pulse oximetry reading) were recorded from the first ED visit within the previous 14 days (the “index visit”). The form also documented presence or absence of 18 home medications and 39 questions about past medical history. Outcomes, including results of repeated molecular testing, or antibody testing for SARs-CoV-2 were recorded up to 30 days after the date of the SARS-CoV-2 test that qualified the patient for enrollment.

The criterion standard for disease positive was evidence of SARs-CoV-2 infection, from either a positive molecular diagnostic test from a swab sample (usually from the nasopharynx), or a positive serological IgM or IgG antibody, documented within 30 days of enrollment. The criterion standard for disease negative required that patients have no positive molecular or serological test for SARS-CoV-2 or clinical diagnosis of COVID-19 within 30 days.

### Model development

After upload, each form was inspected centrally for completeness and sensibility of data, resulting in verification queries. For example, if a patient had none of 28 symptoms, the site investigator was asked to double-check the medical record. Uploaded records were considered eligible for analysis after resolution of queries coupled with electronic verification by the data abstractor and site investigator that each uploaded record was a true and complete reflection of data in the medical record. The *a priori* plan for model development called for a classical approach of screening candidate variables with unadjusted analyses, followed by multivariable logistic regression analysis with conversion into either a scoring system or a set of criteria. The data collection form was produced in March 2020, when the phenotype of patients with SARs-CoV-2 infection was incompletely understood. Thus, the plan was an agnostic approach: to screen all potential predictor variables for discriminative value, including method and day of arrival to the ED, patient demographics, symptoms, vital signs, contact risks, habits, medications and past medical history. As previously described, we estimated a minimum sample size of 20,000 to allow derivation and validation on approximately 10,000 patients in each step, assuming a 30% criterion standard positive rate and a greater than 10:1 ratio of outcomes to predictor variables, recognizing this as a minimum criterion.^9,12,14^

To derive the CORC rule, we first extracted a 50% random sample, and, for statistical testing, used the criterion standard result as the dependent variable. Per protocol, categorical data that were not charted were considered absent, but missing continuous data (>0.1%, age, vital signs, and body mass index) were analyzed for monotonicity and replaced using multiple imputations method in SPSS® (IBM Corp. Released 2020. IBM SPSS Statistics for Windows, Version 27.0. Armonk, NY: IBM Corp). The mean values from five iterations were used, and compared with the pre-imputation mean to confirm a significant change (P<0.05). Bivariate data were compared between test + and test - using the Chi-Square statistic and means from parametric predictors (e.g., age) were compared using unpaired t-test. Variables with P<0.05 were entered into logistic regression equation, initially leaving parametric data as continuous (to create the CORC score), and variables selected for rule development using an empirical stepwise forward approach using the likelihood ratio approach. Model fitness was assessed with the Hosmer-Lemeshow test.

To produce the actual CORC rule (a simplified version of the logistic regression equation), we dichotomized continuous data at the midpoint of the difference in means between patients with and without SARS-CoV-2 infection. To test for validity, the CORC score was computed by solving for probability (P) from the logistic regression equation; the net positive points for the CORC rule were calculated for each of the remaining 50% of patients in the registry, who were independent of the derivation population. Diagnostic accuracy of the CORC score and rule was assessed in the validation with receiver operating characteristic curve and diagnostic indexes from contingency table analysis. Data were analyzed with SPSS® software with the Complex Sampling and Testing module.

## RESULTS

Data for this analysis were downloaded from the registry on December 3, 2020. The download included 20,060 complete records collected per protocol from 41 hospital systems representing 116 unique hospitals from 25 states. Eligible records came from patients tested for SARS-CoV-2 from the first week of February, until the fifth week of October, 2020. After exclusion of 210 records marked by the sites as screen fails (from a later discovered exclusion criterion), 19,850 records were left for analysis.

Multiple imputation successfully replaced all missing parametric values, including body mass index as the most frequently missing value (in 25%), followed by respiratory rate (1.4%). For age, blood pressure, and pulse oximetry, values were missing in less than 1% of the sample. Each record was then assigned a random number drawn from 1 to 19,850, and re-sequenced, and the first and second halves were used to derive and test the CORC score, respectively. **Table 1** shows the clinical characteristics of the sample, divided into the derivation and validation groups, and indicates that random sampling produced two comparable groups. Compared with US Census Bureau data from 2019, this ED sample is older, has a higher representation of persons identifying as Black (and lower percentage of persons identifying as White), but a similar distribution of biological sex and Hispanic or Latino ethnicity.^15^ Table 1 conveys findings that are important to developing accurate pretest probability criteria using criteria available at the bedside. First, the pooled prevalence of infection among those tested was 34%, which is relatively high for producing exclusionary criteria. Second, the mean turn-around-time for SARs-CoV-2 testing was greater than one day, although the median time was 0.5 days (interquartile range 0-1.0). Third, approximately 5% of the sample had none of the 28 recorded symptoms at presentation, but still had clinical suspicion that led to testing. Fourth, approximately one-quarter of all patients had no chest radiograph performed and almost one-quarter had no laboratory analysis of a blood specimen. Additional data of relevance include the fact that 1,915 patients (10%) had visited the ED within the previous 14 days prior to testing for SARS-CoV-2, but records documented clinical suspicion for COVID-19 in only 367 (19%) of these visits.

**Table 1.**
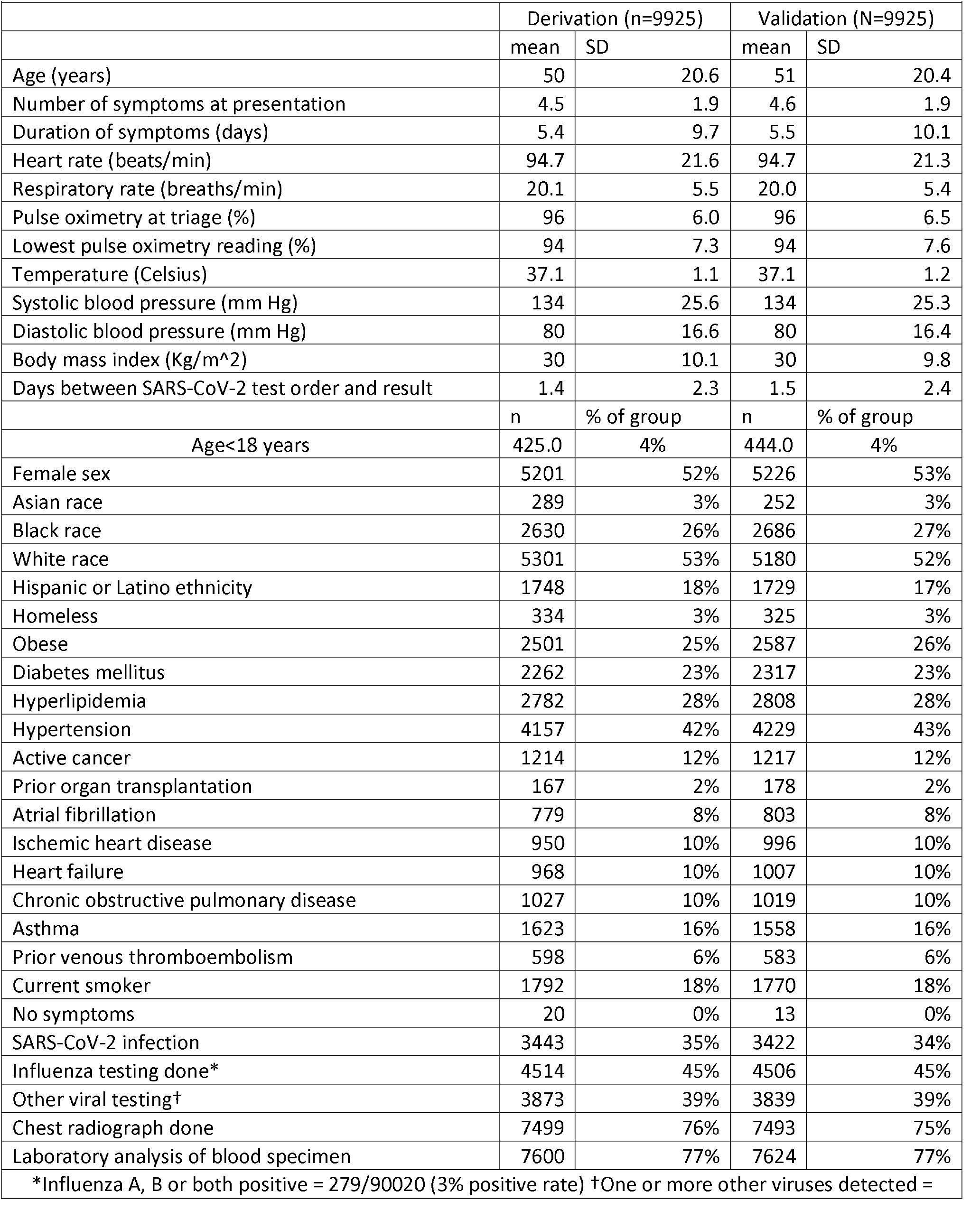
Clinical features of the derivation and validation samples

Of 107 candidate predictors shown in **supplemental Table 1**, 72 had P<0.05 by univariate statistical analysis (Chi-Square for bivariate data and unpaired t-test for continuous data), comparing data from patients with positive SARS-CoV-2 testing versus patients with negative test. These 72 variables were subsequently evaluated by stepwise forward multivariable logistic regression using the likelihood ratio method. After exclusion of 42 variables that were not significant, the procedure was repeated with 30 predictors. The model selected for the CORC score was from step 13 (13 variables) based upon consideration of the need to limit number of predictor variables for practical use with maintenance of model fitness by keeping the Hosmer-Lemeshow P value >0.10. These 13 variables were then examined by a single-step logistic regression to produce **Table 2**. This model produced a C-statistic (area under the receiver operating characteristic curve) of 0.80 (0.79-0.81). When the equation solved for probability, at a cutoff of 0.1 this yielded sensitivity of 97% and specificity of 20% in the derivation population. **Table 3** shows the simplification of the CORC score into the 13 component CORC rule, which included the dichotomization of age, temperature and the pulse oximetry reading obtained at the time of triage in the ED. With the exception of the anosmia/dysgeusia variable, the use of whole digits (−1 or +1, score range 0-13) proportionatly reflect the sign and rounded magnitude of the beta coefficients and intercept obtained from repeated logistic regression with age, pulse oximetry and temperature converted to dichotomous variables with cutoffs at 50 years, 94.5% and 37.5°C respectively. With 0 or fewer points considered a test negative CORC rule result, a negative CORC rule produced diagnostic sensitivity of 96% and specificity of 21% in the derivation population.

**Table 2.** Logistic regression results of the selected model (the CORC score)

|  |  |  |  |  |
| --- | --- | --- | --- | --- |
|  | Coefficient | Odds ratio | 95% CI |  |
|  |  |  | Lower | Upper |
| Black race | 0.88 | 2.40 | 2.04 | 2.82 |
| White race | -0.42 | 0.66 | 0.57 | 0.76 |
| Hispanic or Latino ethnicity | 1.34 | 3.81 | 3.28 | 4.42 |
| Age in years | 0.02 | 1.02 | 1.02 | 1.02 |
| Symptom: Loss of sense of taste or smell | 1.93 | 6.89 | 5.22 | 9.11 |
| Symptom: Non-productive cough | 0.43 | 1.54 | 1.39 | 1.70 |
| Symptom: Fever | 0.44 | 1.55 | 1.39 | 1.72 |
| Symptom: Muscle aches | 0.48 | 1.61 | 1.43 | 1.81 |
| Exposure to COVID-19: None known | -0.45 | 0.64 | 0.57 | 0.71 |
| Exposure to COVID-19: Household contact with known or suspected infection | 1.68 | 5.36 | 4.42 | 6.51 |
| Pulse oximetry at triage | -0.04 | 0.96 | 0.95 | 0.97 |
| Temperature in Celsius | 0.44 | 1.55 | 1.46 | 1.65 |
| Current smoker | -0.81 | 0.45 | 0.39 | 0.51 |
| Intercept | -14.78 | N/A |  |  |
| Model analysis: Hosmer Lemeshow P=0.526, McFadden's pseudo R<sup>2</sup>=0.22, C statistic= 0.80 |  |  |  |  |

**Table 3.** The COVID-19 rule out criteria (CORC rule)

| **Table 3. The COVID-19 rule out criteria (CORC rule)** |  |
| --- | --- |
| Component | Points |
| Black race | +1 |
| White race | -1 |
| Hispanic or Latino ethnicity | +1 |
| Age >50 years | +1 |
| Symptom: Loss of sense of taste or smell | +1 |
| Symptom: Non-productive cough | +1 |
| Symptom: Fever | +1 |
| Symptom: Muscle aches | +1 |
| Exposure to COVID-19: None known | -1 |
| Exposure to COVID-19: Household contact with known or suspected infection | +1 |
| Pulse oximetry at triage <95% | +1 |
| Temperature in Celsius >37.5 | +1 |
| Current smoker | -1 |

When applied to the other half of the sample (validation group, n=9925), the CORC score and rule performed similarly. The CORC score had an area under the receiver operating characteristic of 0.80 (95% CI, 0.79 to 0.81), and when solved for probability (P), if P<0.1, the diagnostic sensitivity was 96.8% (96.1 to 97.3%) and the specificity was 20.1% (19.1 to 21.1%), yielding a posterior probability of 7.8% (6.5 to 9.8%). The accuracy of the CORC score in the validation dataset was maintained across the month of diagnosis. For the 8,444 patients evaluated early in the US pandemic (February to May 2020) the area under the receiver operating characteristic curve for the CORC score was 0.80 (0.79 to 0.81), compared with 0.81 (0.79 to 0.84) for 1,481 patients evaluated from June 2020 onward. The CORC score area under the receiver operating characteristic curve was decreased in patients with zero symptoms (0.73, 0.69 to 0.77).

In the validation set, the CORC rule negative (0 or fewer points from Table 3) produced sensitivity of 95.6% (94.8 to 96.3%), specificity of 20.0% (19.0 to 21.0%), likelihood ratio negative of 0.22 (0.19 to 0.26) and a posterior probability of 10.4% (8.9 to 12.1%). The probability of infection increases with the number of positive points from the CORC rule. This stepwise, positive concordance is shown in **Figure 1**, which plots the posterior probability of positive SARS-CoV-2 testing as a function of the number of points from the CORC rule from the validation population. The probability of SARs-CoV-2 infection is >75% in a patient with +5 or more points from the CORC rule.

**Figure 1.**
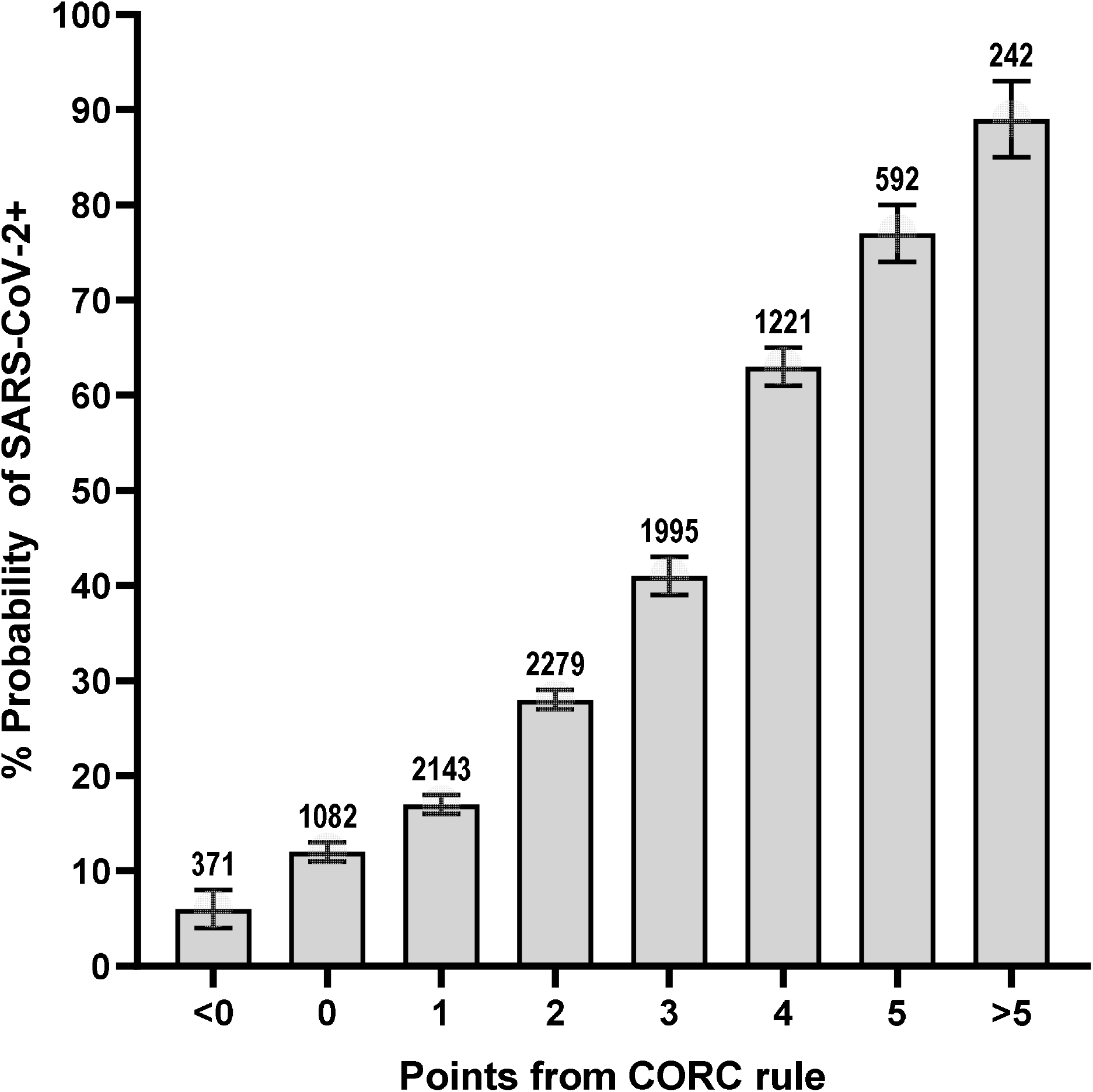
Probability of SARS-CoV-2 infection according to number of points from the CORC rule

Given the concern about low diagnostic sensitivity for molecular testing on swab samples, a relevant question is how the CORC rule performed among patients with an initially negative swab test who had subsequent evidence of SARS-CoV-2 infection within 30 days. From the entire sample (both derivation and validation), the initial swab that qualified the patient for enrollment was negative in 13,159 patients. Of these, 174 (1.1%) subsequently had evidence of SARs-CoV-2 from either a repeated nasopharyngeal swab or positive antibody testing done within 30 days. Among these 174 patients who had a possibly false negative molecular test done on a swab sample, the CORC rule was >0 in 87%.

## DISCUSSION

This work addresses the urgent need for criteria to rapidly, easily, and accurately estimate the probability of SARs-CoV-2 infection. Using registry data from the RECOVER Network, which was specifically created to address this knowledge gap, we found that 13 variables—11 of which were obtained from verbal interview, together with one data point each from a thermometer and a pulse oximeter--can accurately predict the probability of SARs-CoV-2 infection if entered into a logistic regression equation and solved for probability (the CORC score) ^9^. A simpler version comprising 13 binary variables, scored with negative or positive point values (the CORC rule, shown in Table 3) provides similar accuracy.

The CORC score was derived from a large patient pool enrolled from 27 states with demographics reflective of the overall US population. The overall utility and durability of the CORC score is suggested by the area under the receiver operating characteristic curve of 0.80 in both the derivation and validation samples, and that this level of accuracy was retained in patients tested either earlier or later in the first year of the US SARS-CoV-2 pandemic. A negative CORC rule (0 or fewer points) had 95.6% sensitivity and 20.0% specificity in the validation sample, providing a likelihood ratio negative of 0.22 (95% CI 0.19 to 0.26). Moreover, the CORC rule was positive (>0 points) in the 87% of patients with an initially negative and subsequently positive molecular test for SARS-CoV-2 done on a swab sample from the nasopharynx. For the goal of predicting high risk of infection, patients in the validation sample with a +5 or more points from the CORC rule had a >75% probability of a positive test.

In terms of methodological strength, the large, diverse, and representative patient sample of patients tested for SARS-CoV-2 has a low risk of sampling bias, which has hampered previous prediction rules for COVID-19.^16^ The practical benefit of the CORC criteria is the lack of requirement for radiological or laboratory data, which were not ordered in the usual care of over a quarter of patients in this sample and are not available in many settings where risk assessment for probability of SARS-CoV-2 infection is critical to decision-making. These findings suggest that the CORC rule, if validated in prospective work, can assist with decisions about need for formal diagnostic testing and isolation procedures for persons passing through high throughput settings including the triage area of some emergency departments and medical clinics, and at the point of entry for homeless shelters, industry, correctional facilities, and extended care facilities. In the home setting thermometers are common and in some protocols pulse oximetry has been used to monitor outpatients with known COVID-19.^17^ Thus, in concept, the CORC rule could be an adjunctive measure to assess the probability of SARs-CoV-2 among household contacts of persons known to be infected.

The data for the CORC score and rule were obtained retrospectively using rigorous methods to ensure high value predictor variables, and unique, relevant circumstantial data. In contrast to many recent reports using clinical informatics, the level of detail for the data from this study required manual evaluation of medical records by research personnel. For example, manual review was required to ensure that the symptoms recorded represented those that the patient manifested on the first contact with the healthcare system while infected with SARs-CoV-2—which was the case for 1,915, or 10% of the cohort. The CORC score was a required step to create the simpler CORC rule. The more formal CORC score is calculated by exponentiating the logistic regression equation and solving for probability, a task easily performed using an online or internet-based calculator. However, recognizing prior literature on the real-world behavior of physicians, we believe a simpler scoring system comprising positive or negative whole single digits will enhance dissemination and adoption.^18,19^

The predictor variables retained by the selection process for both the CORC score and rule warrant discussion. First, the large sample and high prevalence would have allowed the stepwise forward logistic regression to retain many more variables, but we terminated the selection at 13 variables for several reasons. The first reason centered on the pragmatic consideration of the time required in busy clinical practice to use the decision aid. The second reason is concern about an overfit model, which is more likely to occur with an excessively complex derivation, regardless of the learning method.^20^ Previous simulation studies suggest that the variability of the area under the receiver operating characteristic curve with 13 predictor variables, and a sample size of 10,000 including >30% prevalence of outcomes is less than 5% with repeated sampling.^21^ Third, at the 14^th^ step, the model began to introduce variables that might be more vulnerable to interobserver variability, including “active cancer” at step 14, a predictor that likely requires more inference than the retained 13 variables. A potentially unanticipated finding is that smoking history was retained as a negative predictor of infection—a finding that has been reported by others.^22-24^

Compared with persons identifying as White, Black race and Latino/Hispanic ethnicity significantly increased the probability of infection. Race-specific patterns in symptom manifestation that might alter clinical suspicion and testing threshold do not appear to explain the differences in positive rate.^25^ To our knowledge, no genetic or biologic reasons explain why Black and Hispanic/Latino patients are more likely to have a SARS-CoV-2 infection. Instead, the statistical weight on these variables may result from them acting as proxies for other societal factors. The association of race with positive testing with may correlate with a higher likelihood of working service-related jobs which are unable to be done from home (thereby increasing exposure to SARS-CoV-2). In one study of a cohort of SARS-CoV-2 infected patients in Louisiana, 77% of those requiring hospitalization were Black; only 30% of the total area population is Black.^26^ However, when adjusted for socioeconomic status and pre-existing clinical comorbidities, there was no racial difference identified in mortality.^26^ Ongoing work will report the impact of insurance status and geographic location (by four digit zip code) on SARS-CoV-2 infection rate and severity.

The retrospective collection of data introduces the primary limitation of this work inasmuch as the CORC score and rule performance, including metrics of inter-rater reliability and operational characteristics, have not been used yet in real practice. For example, in terms of generalizability for high throughput screening, it remains unknown whether the temperature component, measured by an infrared thermometer, and the oxygen saturation, measured by a portable pulse oximeter, would provide similar diagnostic accuracy. Based upon the estimated likelihood ratio negative of 0.22, a CORC rule of zero or less would allow a very low posterior probability (e.g., <2.0%) in populations with a relatively low prevalence of infection (<9%).^27^ Symptoms not recorded were assumed to be absent, which could affect rule precision and accuracy. Another limitation is the relative lack of data from most recent cases. The most recent patient was evaluated in October and most cases came from early spring of 2020. The genotype of the virus, as well as the phenotype of infected patients, may have changed with time, and the effect on accuracy and imprecision are unknown. Additionally, it remains possible that machine-based learning methods may offer a superior role, although one study using ED-based data that directly compared three derivation techniques, including backward stepwise logistic regression, decision-tree based approaches random forest, and a gradient boosted decision tree method found logistic regression to produce a higher area under the curve for prediction of infection.^28^ To consider the possible benefit of machine learning, the authors reviewed the diagnostic accuracy of criteria of 20 reports to predict SARS-CoV-2 infection, including both logistic regression and machine learning techniques.^28-30^ This informal scoping review revealed that the diagnostic accuracy of machine learning was not superior to logistic regression-based models, and therefore supported the pre-planned classical approach to model development.^28^

In conclusion, we present novel criteria requiring only information that can be obtained from the patient interview, a thermometer, and a pulse oximeter to predict the probability of SARS-CoV-2 infection. A score of zero from the simplified COVID-19 rule-out criteria (the CORC rule) predicts a low probability of infection and a score of 5 or more predicts a high probability of infection. If prospectively validated, we believe the CORC rule will help expedite decision-making in high throughput settings.

## Data Availability

Available to public after June 2021

## Data Availability

All code used in this project is publicly available here: https://osf.io/p9ntr/ Due to individual site IRB restrictions, data is not publicly available. Please contact the ENIGMA CHR Working Group co-chairs if you are interested in joining and participating in the working group. Then you will be able to submit secondary data proposals.

## Acknowledgments

The authors thank Patti Hogan and Amanda Klimeck for administrative oversight of the RECOVER Network.

